# Sleep timing irregularity in midlife: Association with incident major adverse cardiac events and cardiovascular disease mortality over a 10-year follow-up

**DOI:** 10.1101/2025.11.04.25339506

**Authors:** Laura Nauha, Maisa Niemelä, Saeid Azadifar, Raija Korpelainen, Vahid Farrahi

## Abstract

**Background:** Sleep timing reflects daily routines and lifestyle patterns, which influence cardiovascular health through circadian mechanisms that regulate cardiovascular processes. Wearable devices enable sensor-based assessment of sleep timing, offering insights into daily behavior. This study examined how the regularity of wearable device–determined sleep timing (bedtime, wake-up time, and sleep midpoint) predicts incident major adverse cardiac event (MACE) and cardiovascular disease (CVD) mortality over a 10-year follow-up in midlife.

**Methods:** The study included 3,231 participants (39.5% men) from the Northern Finland Birth Cohort 1966 who attended the 46-year follow-up in 2012–2014. Participants were followed until December 31, 2023, or until a MACE (acute myocardial infarction, unstable angina, stroke, heart failure hospitalization, or CVD death) or were censored due to moving abroad or dying from a non-cardiovascular cause. Sleep timing regularity was assessed via 7-day standard deviation for bedtime, wake-up time, and sleep midpoint, categorized into tertiles: regular, fairly regular, and irregular. Cox proportional hazards models estimated hazard ratios (HRs) with 95% confidence intervals (CIs), adjusting for gender, employment status, body mass index, systolic blood pressure, glycated hemoglobin, low-density lipoprotein cholesterol, and total physical activity. Analyses were stratified by sleep duration below or above the group median (7 h 56 min).

**Results:** In total, 128 participants (4.0%) experienced MACEs during the follow-up period. Irregular sleep timing was associated with an elevated risk, but this association was observed only among participants whose sleep period was shorter than the group median. Individuals with irregular bedtimes had a 2.01-fold higher risk of MACEs compared to those with regular bedtimes (HR = 2.01, 95% CI: 1.00–4.01, p = 0.049), and those with irregular sleep midpoints had a 2.00-fold higher risk compared to those with regular midpoints (HR = 2.00, 95% CI: 1.01–3.98, p = 0.048).

**Conclusions:** Among the participants with sleep durations under eight hours, irregular sleep timing was a significant risk factor for MACEs. Specifically, variability in bedtime and sleep midpoint, but not in wake-up time, was associated with increased risk. These findings highlight the importance of consistent sleep behavior, particularly regular bedtimes, as a potential target for health promotion.

## BACKGROUND

The 24-hour internal circadian rhythm is an important factor in human biology, regulating the circadian rhythms that influence sleep, metabolism, and cardiovascular function (1). Critically, disruptions in circadian alignment, such as those caused by evening chronotype, shift work, irregular sleep timing, and late meal timing, have been associated with adverse cardiometabolic health (2–6). These findings suggest that behavioral patterns affecting circadian stability may play an important role in cardiometabolic health.

Research has shown that any amount of physical activity could be beneficial for cardiovascular health (7,8) and, relatedly, that higher physical activity level is associated with reduced risk of cardiovascular disease (CVD) and mortality (9,10). In addition to physical activity, both short and long sleep durations have been linked to increased risk of CVD (11,12), with adequate sleep duration and quality being considered important in maintaining cardiometabolic health (9,13–15).

Sleep health, however, is multidimensional. In addition to sleep duration and quality, regularity of sleep and sleep timing could also be predictors of health outcomes. Recent studies have increasingly emphasized the role of regularity in sleep and activity behaviors in promoting cardiometabolic health (5,6,16,17). In a recent statement, the American Heart Association emphasized the multidimensionality of sleep health, listing sleep duration, regularity, timing, continuity, satisfaction, daytime functioning, sleep architecture, and the absence of sleep disorders as essential elements contributing to healthy cardiovascular functioning (15).

Emerging research has also highlighted the potential causal relationships between sleep-related factors, such as chronotype, sleep duration, and insomnia symptoms, and cardiometabolic outcomes, including blood pressure, cholesterol levels, and type 2 diabetes (18–20). Inconsistent sleep rhythms and shorter sleep durations have additionally been associated with elevated cardiometabolic mortality risk (5). Indeed, a large-scale longitudinal study from the UK Biobank found that higher night-to-night variation in sleep duration was associated with increased risk of myocardial infarction and stroke, independent of genetic risk (21).

Notably, the increasing availability of wearable devices offers new opportunities to assess sleep and activity behaviors in everyday life with increased temporal resolution and objectivity, enabling more accurate evaluations of these factors’ long-term health implications (22). This advancement has, in turn, facilitated the quantification and computation of different metrics reflecting the regularity of sleep patterns and timing. Device-based metrics, such as the sleep regularity index, interdaily stability, and standard deviation (SD) of sleep timing, have shown consistent associations with cardiometabolic biomarkers, including blood pressure, waist circumference, and lipid profiles (16,23–26). These metrics, though, are not identical, and their related interpretations and implications may differ. First, the sleep regularity index quantifies the probability that an individual is in the same sleep or wake state at any two time points in time 24 hours apart, averaged over multiple days (27). Interdaily stability, meanwhile, measures the stability of rest-activity rhythms across multiple days, indicating how consistent a person’s daily pattern is over time (28). In contrast, SD of sleep timing provides a direct measure of variability in specific behaviors (e.g. bedtime or wake-up time) offering a simple yet informative indicator of sleep routine consistency. While the sleep regularity index and interdaily stability capture general rhythmicity and alignment with the 24-hour cycle, they are composite measures that do not isolate specific behavioral components.

To complement these composite metrics and better understand the role of specific sleep timing behaviors in cardiovascular health, this study focused on sleep timing regularity. By examining the consistency of bedtime, wake-up time, and the midpoint of the sleep period, we aimed to capture direct measures of routine variability that may be more interpretable and actionable in clinical and public health contexts. This study investigated whether sleep timing regularity measures predict incident major adverse cardiac events (MACEs) and CVD mortality over a 10-year follow-up in a middle-aged population from the Northern Finland Birth Cohort 1966. Sleep timing regularity was assessed based on the regularity of bedtime, wake-up time, and the midpoint of the sleep period.

## METHODS

### Study population

The study comprised data from the Northern Finland Birth Cohort (NFBC1966), a longitudinal birth cohort that encompasses all Oulu and Lapland newborns whose births were expected in 1966 (N = 10,331) (29,30). The cohort members were regularly monitored prospectively through a wide range of clinical measurements, interviews, and postal questionnaires.

The present study included those members of the NFBC1966 who participated in the most recent follow-up study at the age of 46 years (between 2012 and 2014), agreed to wear accelerometer-based activity monitors, and had sufficient accelerometer data. The 46-year follow-up included the completion of postal questionnaires on health behaviors (n = 7,146; 69.2%), attending a clinical examination day for the collection of fasting blood samples and anthropometric measurements (n = 5,832; 56.5%), and the two-week 24-hour accelerometer monitoring of daily activity behaviors (n = 5,624; 54.5%). The study was approved by the Ethical Committee of the Northern Ostrobothnia Hospital District in Oulu, Finland (94/2011).

### Measurements

#### Questionnaires and clinical measurements

Participants self-reported their smoking status (non-smoker, former smoker, or current smoker), education level, marital status, and alcohol consumption (calculated to g/day based on responses). Heavy alcohol drinkers were defined according to the alcohol consumption instructions of the Finnish Institute for Health and Welfare, which sets the level for men at ≥ 40 g/day and the level for women at ≥ 20 g/day (31). The shortened morningness–eveningness questionnaire was used to determine the participants’ chronotypes and to assign participants into morning-type, day-type, and evening-type individuals (32,33). Based on the question about their work schedule, the participants were divided into three groups: shift work, day work, and not working or no information available.

The participants attended the clinical examination after fasting for 12 hours overnight and abstaining from smoking and drinking coffee. Trained nurses measured the participants’ heights and weights, and their corresponding body mass indexes (BMI) ([kg/m2]) were calculated. After a period of restful sitting (at least 10 min), their blood pressure was measured using the upper area of each participant’s right arm using an Omron M10-IT automatic blood pressure monitor (Omron M3, Omron Healthcare Europe BV, Netherlands). The recorded blood pressure represented the mean of the three consecutive readings. Participants’ plasma low-density lipoprotein cholesterol was analyzed using fasting blood samples.

#### Device-based measurement of sleep and physical activities

Participants attending the clinical examination day were asked to wear an accelerometer-based activity monitor (specifically Polar Active, Polar Electro Oy, Kempele, Finland) on their nondominant wrist continuously over 24 hours, including while sleeping, for 14 consecutive days (29). The specific monitor used outputs metabolic equivalents (METs) every 30 second using background information (body height, body weight, age, and sex). Polar Active has been shown to better detect daily energy expenditure compared to the doubly labelled water technique (R² = 0.78) (34). Participants did not receive any feedback from the device about their sleep or activity patterns.

#### Sleep period and sleep regularity

Sleep periods were identified from Polar Active MET values using an earlier developed sleep period detection algorithm (16,35). This algorithm identifies bedtime as the start of a prolonged low-activity period and wake-up time as the end of that period and has been shown to estimate bedtime and wake-up time with approximately 20-minute accuracy compared to self-reported sleep diaries and with approximately 5-minute accuracy compared to a smart ring–provided sleep period (35). Notably, smart rings have demonstrated high validity in sleep measurement when evaluated against polysomnography, the clinical gold standard (36).

Sleep regularity was examined from three measures: bedtime (defined as the start of the sleep period), wake-up time (defined as the end of the sleep period), and sleep midpoint (calculated as the halfway point between bedtime and wake-up time). Sleep regularity was quantified using the standard deviation (SD) of bedtime, wake-up time, and sleep midpoint the first 7 consecutive days. A day was considered valid if the non-wear time was less than 2.5 hours, with non-wear time defined as continuous periods with intensity levels below 1 MET. A 7-day period was selected to ensure the studied period included both weekdays and weekend days, allowing for a representative assessment of typical sleep behavior and its variability. This study period aligns with previous research, which has suggested that 5 to 6 nights may be sufficient for a reliable estimation of typical sleep behavior (37).

#### Physical activity

Physical activity during non-sleep periods were calculated using the MET values provided by Polar Active for each day, and the daily means for total physical activity were calculated for each participant. Total physical activity, including all activities with a recorded intensity of 2 METs or higher, was calculated by multiplying each MET value by its duration (total PA [MET min/day]) (33,38). Physical activity intensity-level thresholds were based on the thresholds used by the manufacturer, which have demonstrated better alignment compared to the traditional intensity thresholds originally defined for hip-worn devices (39). For instance, Polar Active’s < 2 MET threshold for sedentary behavior has been found to yield comparable results to the < 100 counts-per-minute criterion used by the ActiGraph GT3X, which is a widely accepted threshold for identifying sedentary time (40).

#### Morbidity and mortality

Data on hospital visits, including discharge diagnoses from inpatient care, were obtained from the Finnish Care Register for Health Care. This nationwide register aggregates information from hospitals operated by municipalities, joint municipal authorities, and the central government, as well as from major private healthcare providers. New MACEs were identified as the first hospital admission or outpatient visit occurring after the conclusion of physical activity monitoring, attributed to any of the following conditions: acute myocardial infarction (ICD-10 codes: I21, I22), unstable angina pectoris (I20), stroke (I63), hospitalization for heart failure (I50), or CVD death (I00–I99). Mortality data, including deaths due to CVD were obtained from Statistics Finland. Incident cases and causes of death were determined based on the primary diagnosis. Participants were followed until December 31, 2023, unless they experienced an earlier MACE or were censored due to moving out of the study area or dying from a non-cardiovascular cause. The study included only participants residing in Finland. Because information on individuals who moved abroad after 2021 was unavailable, censoring for residence outside Finland beyond that point could not be applied. The research data was obtained from Findata, the Finnish Social and Health Data Permit Authority, with data permit THL/6250/14.05.00/2024. Findata was responsible for the pseudonymization of the data and ensuring the anonymity of the final results.

### Statistical Analyses

Only participants with a SD less than 6 hours in each sleep regularity variable were included; higher variability was excluded as outliers. Bedtime, wake-up time, and midpoint regularities were recoded to three equally sized groups and labelled as tertiles: regular, fairly regular, and irregular.

The distributions of sleep-related, sociodemographic, lifestyle, and cardiovascular risk factors were calculated separately for participants with and without a MACE during the 10-year follow-up. The statistical significance of the group differences was analyzed using chi-square tests for normally distributed data and Mann-Whitney U tests for skewed data. For categorical variables, group differences were assessed using chi-square tests. Post-hoc comparisons were adjusted using the Bonferroni correction.

Cox proportional hazards models were conducted to estimate the hazard ratios (HRs) of MACE incidence with 95% confidence intervals (CIs), with these models being created separately for bedtime regularity, wake-up time regularity, and midpoint regularity. Based on previous studies, this study considered the following variables as covariates: total physical activity (MET min/day), sedentary time (min/day), gender (male/female), alcohol use, smoking status, employment status (employed/unemployed), low-density lipoprotein (LDL) cholesterol (mmol/L), BMI (kg/m²), glycated hemoglobin (mmol/mol), systolic blood pressure (mmHg), chronotype (morning, day, or evening type), and sleep-period duration (41–44). Covariates that proved to be significantly associated with survival (gender, employment status, BMI, systolic blood pressure, LDL cholesterol, and total physical activity) were included in the final models.

The proportional hazards assumption was assessed visually and with scaled Schoenfeld residuals. Sleep period did not meet the proportional hazards assumption. Therefore, Cox models were stratified by sleep period duration. Separate models were constructed for participants with sleep period duration at or below the group median (7 h 56 min) and those above it, referred to as the below-median and above-median sleeper groups. Stratification by the median was chosen because it approximates the recommended sleep duration. More groupings were not possible, as the number of events per group was low. Due to the violation of the proportional hazards assumption, employment status was included as a stratification variable in the Cox model. For total physical activity, a time-varying coefficient model using the tt() transformation in the Cox regression (45) was applied, as the proportional hazards assumption was not met.

Kaplan–Meier survival curves were generated separately for the below-median and above-median sleeper groups to examine the association between sleep regularity and survival. For each group, separate curves were plotted for strata of regularity (regular, fairly regular, and irregular) across three variables: bedtime regularity, wake-up time regularity, and midpoint regularity.

Sensitivity analyses were performed to test the stability of the main findings. First, participants who experienced a MACE within the first 2 years of follow-up were excluded to reduce potential reverse causality. Second, shift workers were excluded to account for the confounding effect of irregular work schedules. In both cases, fully adjusted Cox models were re-estimated using the same covariates as in the main analysis.

All data were analyzed using R version 4.3.1.(46). Survival analyses were performed with the survival package, version 3.8.3, and survival plots were created with the R survminer package, version 0.5.0. Statistical significance was set at p < 0.05.

## RESULTS

The final study sample included 3,231 middle-aged participants whose sleep regularity was measured over 7 consecutive nights, resulting in a total of 22,617 recorded nights. During the over-10-year follow-up, 128 participants experienced a MACE. The average follow-up time was 130.1 (interquartile range (IQR) of 10.8) months.

Table 1 presents the main characteristics of the study population. Covariates that indicated association with MACEs included gender (57.0% of men vs. 38.8% of females), employment status (25.8% of those unemployed vs. 17.7% of those employed). Among continuous variables, participants who experienced a MACE had higher median BMI (28.2 kg/m^2^ vs. 25.7 kg/m^2^), systolic blood pressure (131.0 mmHg vs. 123.0 mmHg), glycated hemoglobin (5.6 mmol/mol vs. 5.4 mmol/mol), and LDL cholesterol levels (3.7 mmol/L vs. 3.3 mmol/L) compared to those without a MACE. Statistically significant differences in BMI and systolic blood pressure were observed between participants included in the final analyses and those excluded (n = 393) due to missing data in one or multiple covariates or a prior MACE (p = 0.024; Mann-Whitney U test) but not in other covariates (all p > 0.05; Mann-Whitney U and chi-square tests, data not shown). There were no statistically significant differences in median bedtime, wake-up time, sleep midpoint, or time in bed between participants who experienced a MACE and those who did not. Similarly, the distribution of sleep timing regularity categories did not differ significantly between the groups.

**Table 1.**
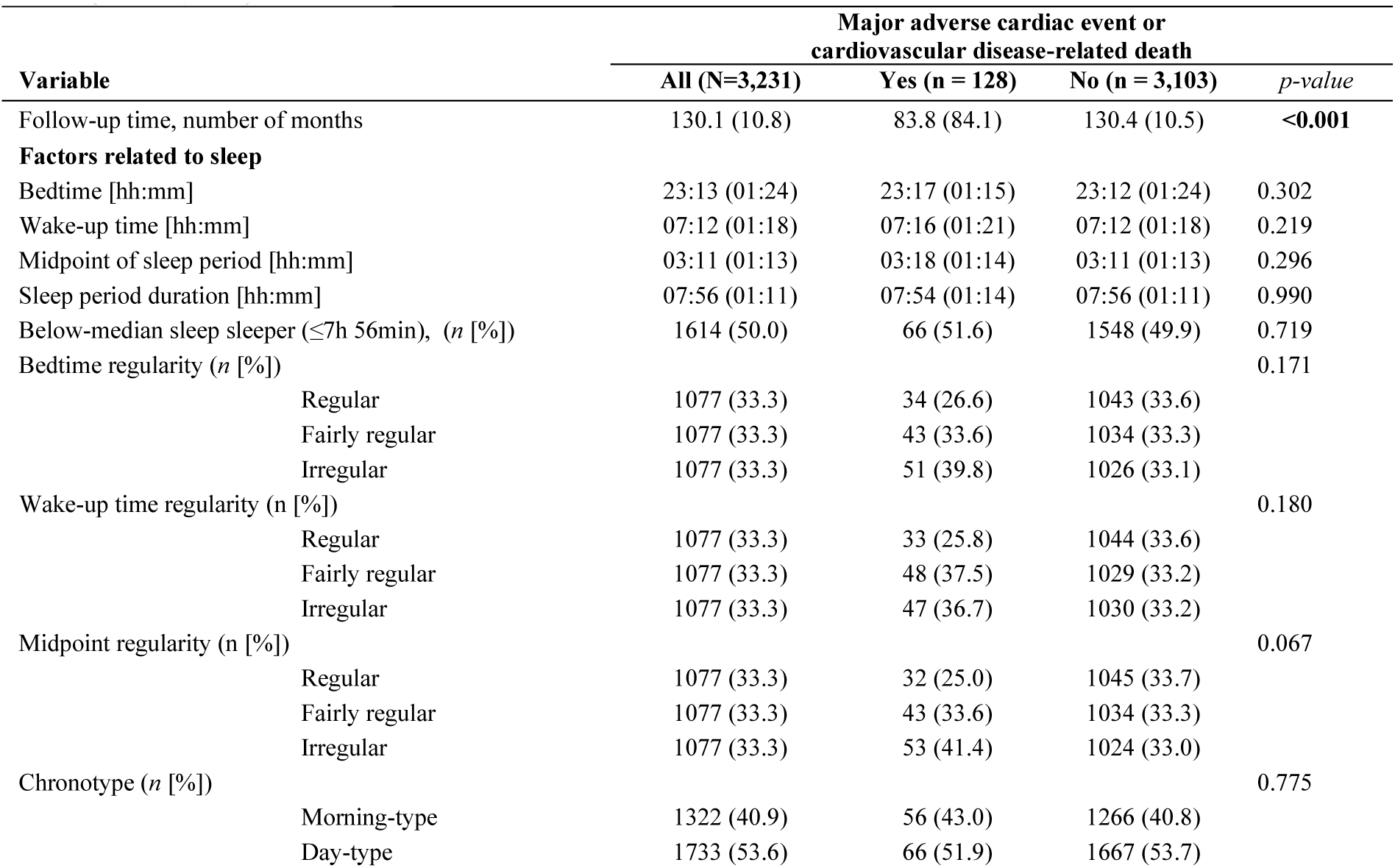
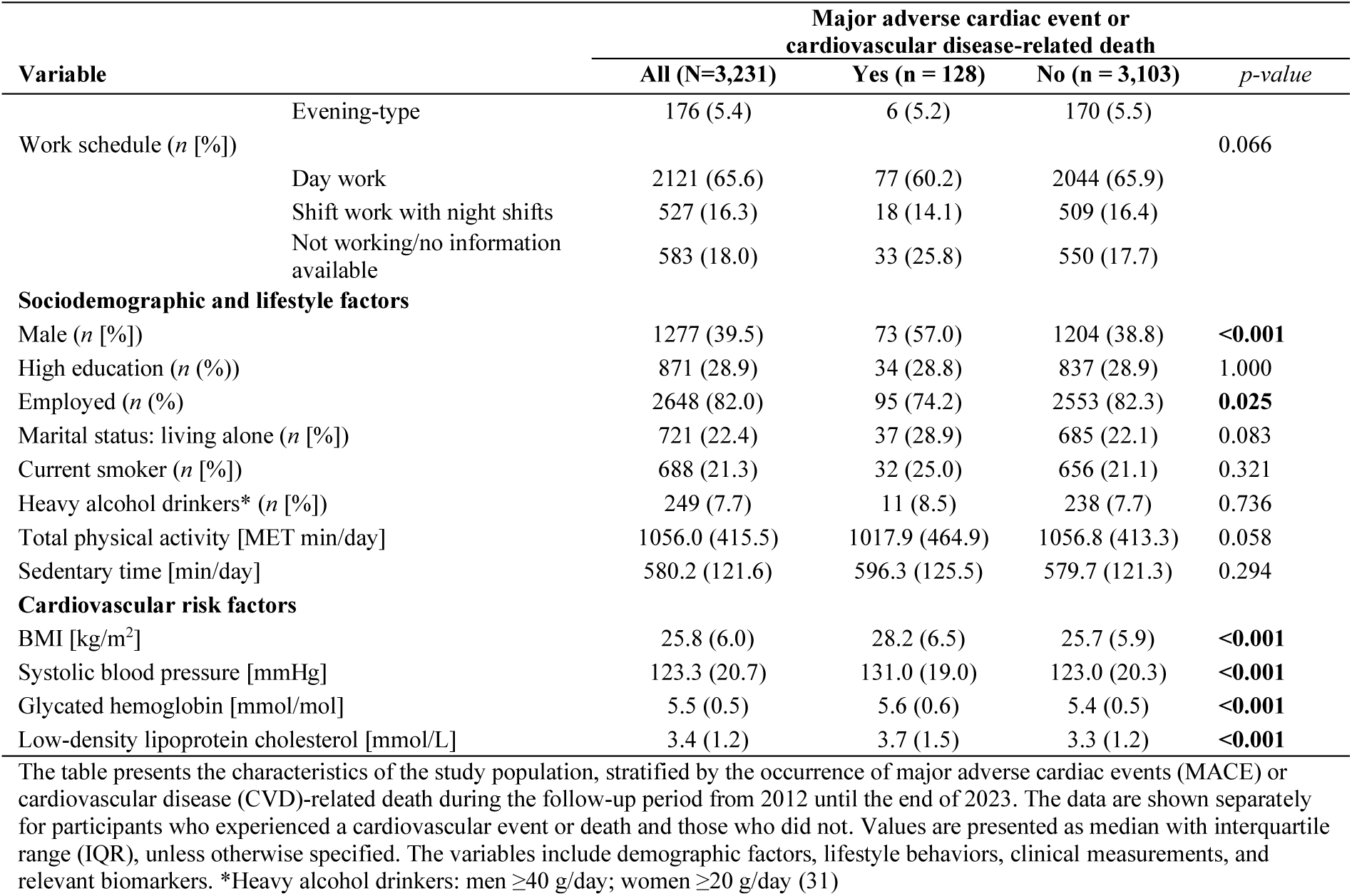
Characteristics of the study population of the Northern Finland Birth Cohort 1966 (N = 3,231)

Median variability values for sleep timing across regularity groups were as follows (Table 1). For bedtime regularity, the median variability was 33 min (IQR 13) in the regular group, 59 min (IQR 15) in the fairly regular group, and 108 min (IQR 55) in the irregular group. For wake-up time regularity, the corresponding values were 41 min (IQR 19), 73 min (IQR 15), and 114 min (IQR 41), respectively. For the sleep midpoint regularity, the median variability was 33 min (IQR 13) in the regular group, 55 min (IQR 11) in the fairly regular group, and 93 min (IQR 40) in the irregular group.

Table 2 presents HRs with 95% CIs, standard errors, and p-values for MACEs in relation to sleep period regularity variables (bedtime, wake-up time, and midpoint regularity). In participants with the below-median sleep periods (≤ 7h 56min), irregular bedtime regularity was associated with an increased risk of MACEs compared to regular bedtime (adjusted HR = 2.01, 95% CI: 1.00–4.01, p = 0.049). Similarly, irregular midpoint regularity was linked to a higher risk of experiencing a MACE (adjusted HR = 2.00, 95% CI: 1.01–3.98, p = 0.048). No significant associations were observed for wake-up time regularity or among participants in the above-median sleeper group (> 7h 56min). The results are also presented as forest plots in Figure 1, which visualizes the HRs and 95% CIs across categories of regularity based on both univariate and adjusted Cox models. The figure highlights the increased risk associated with irregular bedtime and sleep midpoint compared to the reference group, observed only among participants in the below-median sleeper group (≤ 7h 56min).

**Figure 1.**
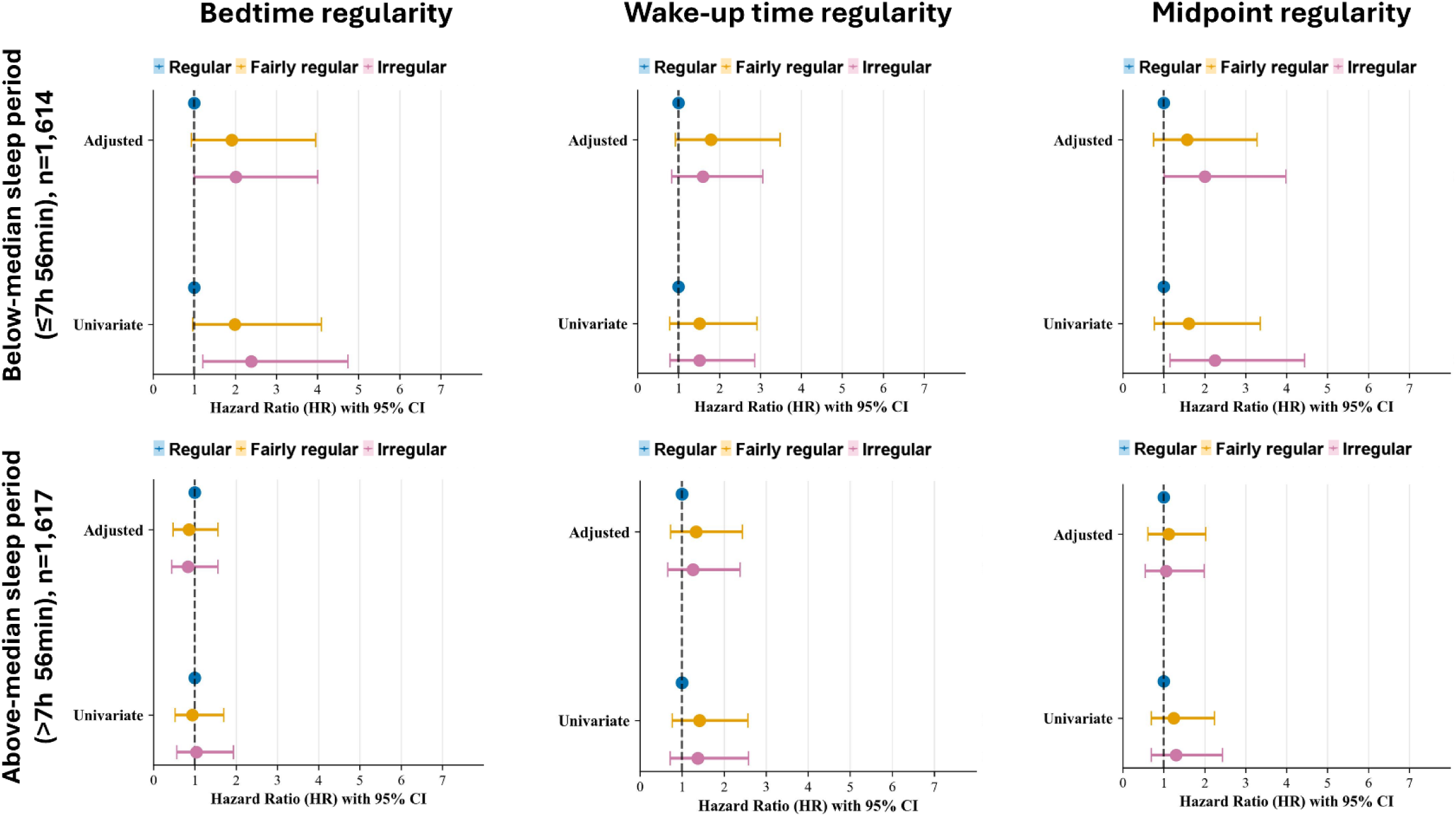
Hazard ratios for sleep regularity by sleep period duration. Forest plots showing hazard ratios (95% CI) for sleep regularity categories, with “regular” serving as the reference group. Results are presented separately for participants with the below-median sleep periods (≤ 7h 56min) and with the above-median sleep periods (> 7h 56min). Adjusted models included the following covariates: gender, employment status, BMI, systolic blood pressure, low-density lipoprotein cholesterol, and total physical activity. Data came from the Northern Finland Birth Cohort 1966 (N = 3,231).

**Table 2.** Hazard ratios for major adverse cardiac events by sleep regularity and sleep period duration.

| Variable | Bedtime regularity |  |  | Wake-up time regularity |  |  | Midpoint regularity |  |  |
| --- | --- | --- | --- | --- | --- | --- | --- | --- | --- |
|  | HR (95% CI) | SE | *p-value* | HR (95% CI) | SE | *p-value* | HR (95% CI) | SE | *p-value* |
| **Below-median sleep period ($\leq 7$h 56min), n = 1,614** |  |  |  |  |  |  |  |  |  |
| **Univariate model** |  |  |  |  |  |  |  |  |  |
| Fairly regular vs. Regular | 1.99 (0.96–4.10) | 0.37 | 0.063 | 1.51 (0.78–2.92) | 0.34 | 0.221 | 1.61 (0.77–3.36) | 0.38 | 0.205 |
| Irregular vs. Regular | **2.39 (1.21–4.74)** | **0.35** | **0.012** | 1.51 (0.79–2.86) | 0.33 | 0.211 | **2.25 (1.15–4.44)** | **0.34** | **0.019** |
| **Adjusted model** |  |  |  |  |  |  |  |  |  |
| Fairly regular vs. Regular | 1.91 (0.93–3.96) | 0.37 | 0.079 | 1.79 (0.92–3.48) | 0.34 | 0.088 | 1.57 (0.75–3.28) | 0.38 | 0.231 |
| Irregular vs. Regular | **2.01 (1.00–4.01)** | **0.35** | **0.049** | 1.59 (0.83–3.06) | 0.33 | 0.163 | **2.00 (1.01–3.98)** | **0.35** | **0.048** |
| **Above-median sleep period ($> 7$h 56min), n = 1,617** |  |  |  |  |  |  |  |  |  |
| **Univariate model** |  |  |  |  |  |  |  |  |  |
| Fairly regular vs. Regular | 0.94 (0.52–1.70) | 0.30 | 0.838 | 1.41 (0.77–2.57) | 0.31 | 0.266 | 1.24 (0.69–2.24) | 0.30 | 0.478 |
| Irregular vs. Regular | 1.04 (0.56–1.94) | 0.31 | 0.890 | 1.37 (0.72–2.58) | 0.32 | 0.334 | 1.30 (0.69–2.44) | 0.32 | 0.412 |
| **Adjusted model** |  |  |  |  |  |  |  |  |  |
| Fairly regular vs. Regular | 0.86 (0.47–1.56) | 0.30 | 0.623 | 1.33 (0.73–2.44) | 0.31 | 0.352 | 1.12 (0.61–2.03) | 0.31 | 0.719 |
| Irregular vs. Regular | 0.83 (0.44–1.56) | 0.33 | 0.555 | 1.26 (0.66–2.38) | 0.33 | 0.487 | 1.05 (0.55–1.99) | 0.33 | 0.884 |

Hazard ratios (HR) with 95% confidence intervals (CIs), standard errors (SEs), and p-values are presented for major adverse cardiac events and cardiovascular disease mortality in relation to 3 sleep period regularity variables: bedtime regularity, wake-up time regularity, and midpoint regularity. Analyses were conducted separately for participants with sleep period durations equal to or below and above the median value of 7h 56min (N = 3,231). Significant associations at the level of $p < 0.05$ are shown in bold. The model was adjusted for gender (male/female), employment status (employed/unemployed), body mass index ( $\text{kg/m}^2$ ), systolic blood pressure (mmHg), low-density lipoprotein cholesterol (mmol/L), glycated hemoglobin (mmol/mol), and total physical activity (MET min/day). Abbreviations: HR = hazard ratio; SE = standard error; CI = confidence interval.

The results of the sensitivity analyses supported the robustness of our findings. First, excluding 12 participants with early MACE events within the first 2 years yielded results similar to those observed in the full sample (data not shown). Second, excluding 527 shift workers, it yielded results comparable to those observed in the full sample. In the below-median sleeper group, irregular bedtime was associated with a higher risk of a MACE compared to regular bedtime (adjusted HR = 2.29, 95% CI: 1.02–5.12, p = 0.044). Likewise, greater variability in sleep midpoint was linked to increased risk of a MACE (adjusted HR = 2.25, 95% CI: 1.01–5.01, p = 0.047). (Table 3).

**Table 3.**
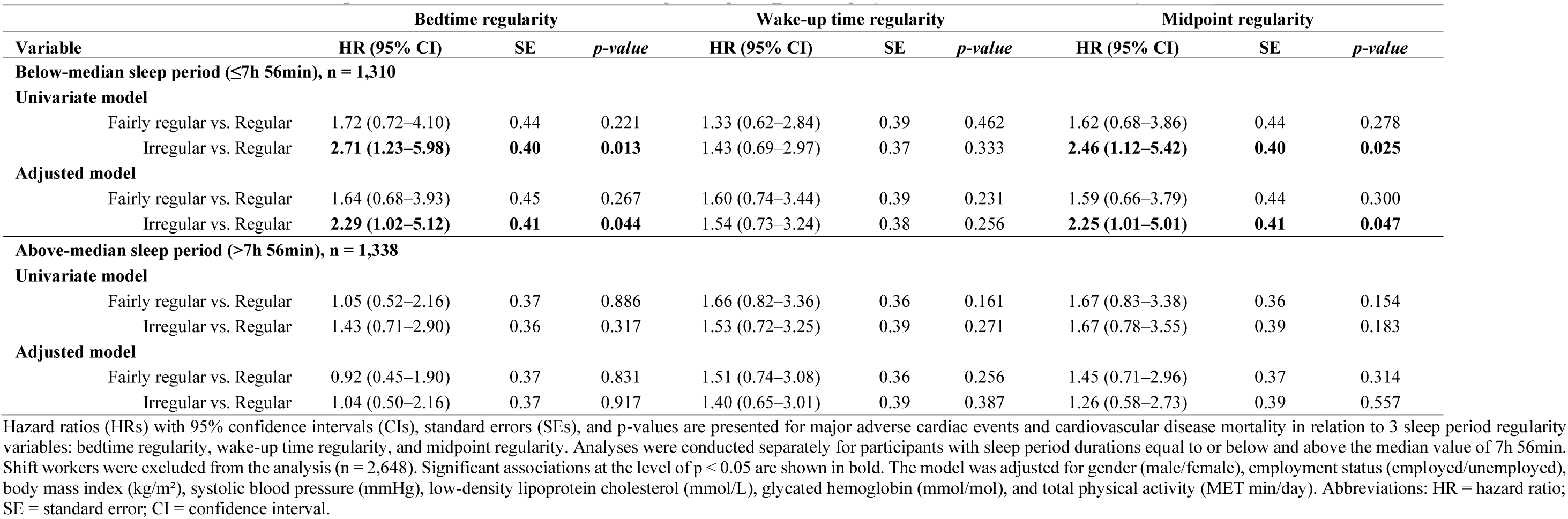
Hazard ratios for major adverse cardiac events by sleep regularity (shift workers excluded)

| Variable | Bedtime regularity |  |  | Wake-up time regularity |  |  | Midpoint regularity |  |  |
| --- | --- | --- | --- | --- | --- | --- | --- | --- | --- |
|  | HR (95% CI) | SE | *p-value* | HR (95% CI) | SE | *p-value* | HR (95% CI) | SE | *p-value* |
| **Below-median sleep period ($\leq 7$h 56min), n = 1,310** |  |  |  |  |  |  |  |  |  |
| **Univariate model** |  |  |  |  |  |  |  |  |  |
| Fairly regular vs. Regular | 1.72 (0.72–4.10) | 0.44 | 0.221 | 1.33 (0.62–2.84) | 0.39 | 0.462 | 1.62 (0.68–3.86) | 0.44 | 0.278 |
| Irregular vs. Regular | **2.71 (1.23–5.98)** | **0.40** | **0.013** | 1.43 (0.69–2.97) | 0.37 | 0.333 | **2.46 (1.12–5.42)** | **0.40** | **0.025** |
| **Adjusted model** |  |  |  |  |  |  |  |  |  |
| Fairly regular vs. Regular | 1.64 (0.68–3.93) | 0.45 | 0.267 | 1.60 (0.74–3.44) | 0.39 | 0.231 | 1.59 (0.66–3.79) | 0.44 | 0.300 |
| Irregular vs. Regular | **2.29 (1.02–5.12)** | **0.41** | **0.044** | 1.54 (0.73–3.24) | 0.38 | 0.256 | **2.25 (1.01–5.01)** | **0.41** | **0.047** |
| **Above-median sleep period ($> 7$h 56min), n = 1,338** |  |  |  |  |  |  |  |  |  |
| **Univariate model** |  |  |  |  |  |  |  |  |  |
| Fairly regular vs. Regular | 1.05 (0.52–2.16) | 0.37 | 0.886 | 1.66 (0.82–3.36) | 0.36 | 0.161 | 1.67 (0.83–3.38) | 0.36 | 0.154 |
| Irregular vs. Regular | 1.43 (0.71–2.90) | 0.36 | 0.317 | 1.53 (0.72–3.25) | 0.39 | 0.271 | 1.67 (0.78–3.55) | 0.39 | 0.183 |
| **Adjusted model** |  |  |  |  |  |  |  |  |  |
| Fairly regular vs. Regular | 0.92 (0.45–1.90) | 0.37 | 0.831 | 1.51 (0.74–3.08) | 0.36 | 0.256 | 1.45 (0.71–2.96) | 0.37 | 0.314 |
| Irregular vs. Regular | 1.04 (0.50–2.16) | 0.37 | 0.917 | 1.40 (0.65–3.01) | 0.39 | 0.387 | 1.26 (0.58–2.73) | 0.39 | 0.557 |

Hazard ratios (HRs) with 95% confidence intervals (CIs), standard errors (SEs), and p-values are presented for major adverse cardiac events and cardiovascular disease mortality in relation to 3 sleep period regularity variables: bedtime regularity, wake-up time regularity, and midpoint regularity. Analyses were conducted separately for participants with sleep period durations equal to or below and above the median value of 7h 56min. Shift workers were excluded from the analysis (n = 2,648). Significant associations at the level of $p < 0.05$ are shown in bold. The model was adjusted for gender (male/female), employment status (employed/unemployed), body mass index (kg/m<sup>2</sup>), systolic blood pressure (mmHg), low-density lipoprotein cholesterol (mmol/L), glycated hemoglobin (mmol/mol), and total physical activity (MET min/day). Abbreviations: HR = hazard ratio; SE = standard error; CI = confidence interval.

Figure 2 shows individual Kaplan–Meier survival curves for the below-median and above-median sleeper groups to examine the association between sleep regularity and survival. Among below-median sleepers (≤ 7h 56min), significant group differences were observed in bedtime regularity (p = 0.036) and midpoint regularity (p=0.048). In both cases, the survival curve for the irregular group declined earlier and more steeply compared to the regular and fairly regular groups.

**Figure 2.**
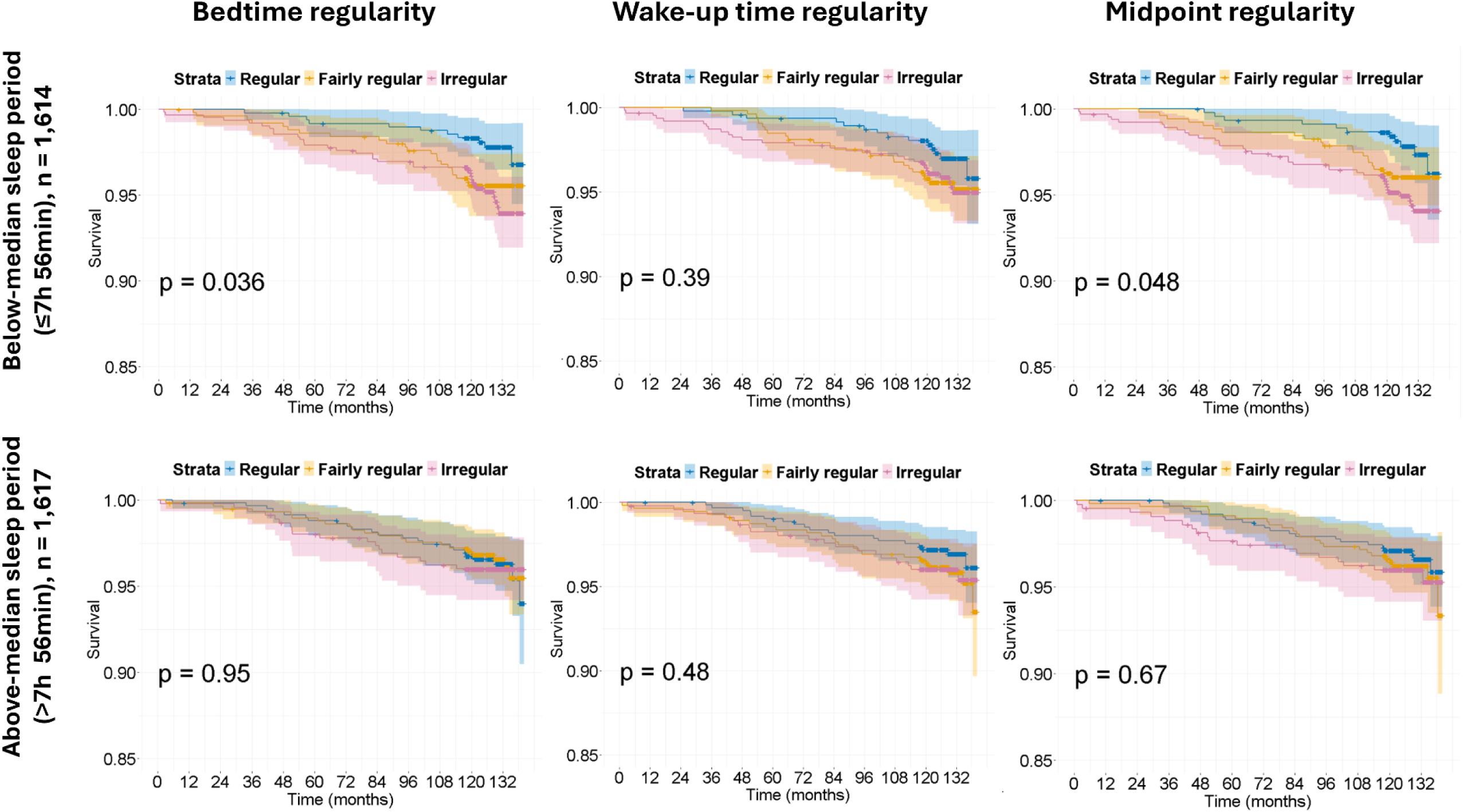
Kaplan–Meier survival curves by sleep regularity and sleep duration. Kaplan–Meier survival curves with p-values and number-at-risk tables are shown, stratified by sleep regularity tertiles, categorized as regular, fairly regular, and irregular. Figures are presented separately for participants with the below-median sleep periods (≤ 7h 56min) and the above-median sleep periods (> 7h 56min). Data came from the Northern Finland Birth Cohort 1966 (N = 3,231), with sleep regularity assessed at midlife. Participants were followed for over 10 years for major adverse cardiac events and cardiovascular disease mortality. Adjusted models included the following covariates: gender, employment status, BMI, systolic blood pressure, low-density lipoprotein cholesterol, and total physical activity.

## DISCUSSION

This population-based study examined the relationship between sleep timing regularity and the risk of MACEs over a 10-year follow-up period among 3,231 middle-aged adults. Average sleep timing and duration were not statistically different between those who experienced an event and those who did not. Conversely, irregular sleep patterns were associated with increased cardiovascular risk among participants with below-median sleep periods (≤ 7h 56min). In this subgroup, individuals with irregular bedtimes or sleep midpoints had a twofold higher risk of experiencing a MACE compared to those with regular bedtimes or regular sleep midpoints. These associations remained statistically significant even after adjusting for other known CVD risk factors, such as BMI, systolic blood pressure, cholesterol levels, glycated hemoglobin levels, employment status, and physical activity.

Our study differs from previous research examining the association between sleep timing regularity and MACEs in a crucial way: namely, in how sleep regularity was operationalized. Similar to our study, previous studies have often used accelerometer-measured sleep, but these prior studies used composite indices like the Sleep Regularity Index (5,26,47,48) or sleep duration variability to operationalize sleep regularity (21). Some studies have further included sleep onset variability with sleep duration variability (17,24). While the findings of those studies have collectively highlighted the importance of sleep regularity, they have not specifically emphasized the full set of behaviorally interpretable sleep variables, such as the regularity of initiating and ending one’s sleep period, which may be more actionable from a public health and behavioral intervention perspective. Our study further contributes to this field through its examination of bedtime regularity, wake-up time regularity, and midpoint regularity as distinct behavioral dimensions of sleep timing.

Notably, higher variability in bedtime and sleep midpoint was linked to increased MACE risk among short sleepers, consistent with prior research showing that irregular sleep onset timing and duration is linked with elevated cardiovascular risk (24). The twofold risk observed in our study between participants with regular and irregular bedtime variability was comparable to previous findings based on a 7-day accelerometer measurement period, where sleep onset variability > 90 minutes was associated with more than double the risk of experiencing a CVD event compared to a variability of ≤ 30 minutes over a median follow-up of 4.9 years. Similarly, although derived using a different methodological approach, our findings align with results of previous studies computing the Sleep Regularity Index from wearable accelerometer data, with these studies also demonstrating that higher sleep irregularity is associated with increased risk of a MACE over a mean follow-up of 7.8 years (47).

On the other hand, wake-up time variability was not associated with MACE incidence in participants with either below- or above-than-median sleep period durations. This finding suggests that wake-up time regularity, unlike bedtime or sleep midpoint, may not be a predictor of cardiovascular events. This finding is noteworthy: Despite growing interest in sleep regularity and its health implications, recent large-scale investigations have not specifically examined whether regularity in wake-up time is associated with MACE incidence (49,50). However, previous finding have suggested that individuals who report waking up either very early (≤ 5 AM) or late (> 7 AM) may have a higher prevalence of hypertension compared to those whose wake-up times fall between 5 AM and 6 AM (51). Similar to the findings produced herein, a population study using questionnaire data did not observe a significant link between wake-up time and cardiovascular mortality, although late bedtimes and delayed sleep midpoint were associated with higher mortality risk (50).

Although there is a scarcity of research examining the associations of wake-up time with CVD risk, several large-scale studies have observed that both earlier and later bedtimes (typically defined as before 10 PM or after midnight) may be associated with modestly elevated risks of all-cause and cardiovascular mortality, particularly when compared to bedtimes between 10 PM and midnight (50–52). A delayed sleep midpoint (typically defined after 3 AM or 4 AM) has also been linked to increased all-cause mortality (50) and hypertension risk (51). Our findings on timing regularity align with these associations with bedtime and midpoint, indicating that both timing and consistency may contribute to cardiovascular risk. As the human circadian system regulates sleep within a 24-hour rest-activity cycle (1), it is possible that maintaining consistent sleep-onset timing may be more critical for supporting sleep quality and cardiovascular recovery than consistent wake-up time alone. Future research should clarify how both the regularity and timing of bedtime and wake-up time contribute to cardiovascular risk.

No significant associations were observed between sleep timing regularity and MACE incidence among participants with sleep period durations above the sample median. This finding contrasts with that of a recent prospective study using device-measured sleep data, which found that irregular sleep was associated with elevated MACE risk, even among individuals who met sleep duration recommendations. Specifically, sufficient sleep duration did not offset the increased risk among irregular sleepers, whereas moderately irregular sleepers appeared to benefit from longer sleep (47). A recent study from the Multi-Ethnic Study of Atherosclerosis further supported this finding, showing that objectively measured regular sleep and sufficient duration were associated with a 39%–42% lower mortality hazard compared with irregular and insufficient sleep, independent of sleep disorders and comorbidities (6). These findings suggest that sleep regularity may play a more critical role than sleep duration in cardiovascular health, possibly due to its influence on circadian alignment and physiological recovery processes (53). In addition, these findings support the view that adequate sleep is a vital component of healthy sleep behavior and may contribute to physiological recovery and resilience against the adverse effects of circadian irregularity. Nonetheless, further research is needed to clarify these mechanisms and their clinical relevance.

After excluding shift workers, we observed an even more pronounced detrimental association between irregular bedtime and sleep periods and the resulting outcomes. This finding suggests that shift work is not the sole factor shaping sleep behavior. In the present study, a notably high proportion of participants who had MACE events or CVD-related deaths (24.4%) were unemployed or had missing employment data compared to the non-MACE group (17.7%). Relatedly, a recent study has shown that unemployment is independently associated with increased cardiovascular mortality, particularly among vulnerable subgroups such as middle-aged women and the elderly (54). In particular, associated mental health challenges may contribute to irregular sleep behavior, which in turn can impact cardiovascular health. Indeed, this relationship is somewhat cyclical: Insomnia increases the risk of depression and anxiety, while psychiatric conditions can disrupt sleep patterns (55). These findings underscore the need to consider broader psychosocial factors, such as unemployment, stress, mental health, and their interactions in understanding the links between sleep behavior regularity and cardiometabolic health.

A major strength of this large-scale population-based study is its comprehensive assessment of sleep timing regularity, including bedtime, wake-up time, and sleep midpoint, in addition to daytime activity behaviors. Another strength of this study stems from its use of comprehensive national health register data, which enabled accurate follow-up regarding cardiovascular events. Unlike many previous studies that have focused specifically on shift workers, our study demographic represents the general middle-aged population across a wide range of socioeconomic and occupational backgrounds, and all participants followed a standardized study protocol. Additionally, despite the length of the study period, moving abroad within the first years of the study proved to be only a minor issue, with only 4 participants doing so. To our knowledge, this is the first population-based study of middle-aged people to study behavioral sleep regularity among a middle-aged population to incorporate an over-10-year follow-up.

This study also has some limitations. The study sample was homogenous regarding participants’ age and ethnicity, which may limit the generalizability of the findings to more diverse populations with different sociodemographic or cultural sleep patterns. Additionally, the methodological limitations of accelerometer-estimated sleep schedules should be acknowledged. Sleep regularity was assessed based on a 7-day consecutive measurement period, which could be considered a relatively short time frame for capturing habitual sleep behavior—though, as mentioned above, previous research has indicated that five to six nights may be sufficient for reliable estimation of habitual sleep (37). While the mean follow-up duration in this study was 10.8 years, extending the follow-up could increase the number of observed cases. Lastly, due to the potential U-shaped association between sleep duration and cardiovascular outcomes (56), a categorization into < 7 h, 7–9 h (recommended), and > 9 h would have been appropriate (57), but we did not apply this due to the small number of events, which may have limited our ability to fully explore the role of sleep duration in these associations.

## CONCLUSION

Our findings suggest an association between the variability in bedtime and sleep midpoint and cardiovascular risk, particularly among individuals with below the sample median sleep period duration which was approximately 8 hours. Of note, irregular sleep timing was not associated with survival risk when the sleep period exceeded 8 hours, indicating that sufficient sleep may offer partial protection regardless of the regularity of sleep timing. Taken together, these results emphasize that consistent sleep behavior, particularly regular bedtimes, should be considered in health promotion to reduce cardiovascular risks.

## Data Availability

NFBC data is available from the University of Oulu, Infrastructure for Population Studies.
Permission to use the data can be applied for research purposes via electronic material request portal. In the use of data, we follow the EU general data protection regulation (679/2016) and Finnish Data Protection Act. The use of personal data is based on cohort participant's written
informed consent at his/her latest follow-up study, which may cause limitations to its use. Please, contact NFBC project center (NFBCprojectcenter(at)oulu.fi) and visit the cohort website for more information.

## LIST OF ABBREVIATIONS

BMI: body mass index
CVD: cardiovascular disease
HR: hazard ratio
MACE: major adverse cardiac event SE standard error
SD: standard deviation

## DECLARATIONS

### Ethics approval and consent to participate

The 46-year follow-up study was approved by the Ethical Committee of the Northern Ostrobothnia Hospital District in Oulu, Finland (94/2011). The participants and their parents provided written consent for the 1966 study.

### Consent for publication

The study was registered in the process of Northern Finland Birth Cohort in 2015. Consent for publication was given on 22 October 2025.

### Availability of data and materials

NFBC data is available from the University of Oulu, Infrastructure for Population Studies. Permission to use the data can be applied for research purposes via electronic material request portal. In the use of data, we follow the EU general data protection regulation (679/2016) and Finnish Data Protection Act. The use of personal data is based on cohort participant’s written informed consent at his/her latest follow-up study, which may cause limitations to its use. Please, contact NFBC project center (NFBCprojectcenter(at)oulu.fi) and visit the cohort website for more information.

### Authors’ Contributions

LN, MN, RK, and VF participated in the literature search; LN, MN, and VF designed the study; LN, MN, RK, and VF collected the data; LN and MN collected the MACE events and information of CVD related deaths from Finnish registers; LN performed the data analysis; SA performed forest plot figures; LN, MN, RK, and VF interpreted the data; and LN, MN, RK, and VF wrote this manuscript. All the authors read and approved the final manuscript.

## Acknowledgments

We thank all cohort members and researchers who participated in the 46-year follow-up study. We also wish to acknowledge the work of the NFBC project center.

